# “You’re just there, alone in your room with your thoughts” A qualitative study about the impact of lockdown among young people during the COVID-19 pandemic

**DOI:** 10.1101/2021.04.11.21254776

**Authors:** Alison R. McKinlay, Tom May, Joanna Dawes, Daisy Fancourt, Alexandra Burton

## Abstract

**Background:** Adolescents and young adults have been greatly affected by quarantine measures during the coronavirus-19 pandemic. Quantitative evidence suggests that many young people have struggled with their mental health throughout “lockdown”, but little is understood about the qualitative impact of social distancing restrictions on mental health, wellbeing and social life. We therefore sought to elicit the views and experiences of adolescents and young adults living in the UK during the pandemic.

**Methods:** Semi-structured qualitative interviews were undertaken with 37 participants aged 13-24.

**Results:** We identified 4 superordinate themes most commonly described by participants about their experiences during the pandemic, including: a) missing social contact during lockdown, b) disruption to education, c) changes to social relationships, and d) improved wellbeing during lockdown. Although we identified some positive experiences during the pandemic, including an increased awareness of mental health and stronger relationship ties, many said they struggled with loneliness, a decline in mental health, and anxiety about socialising after the pandemic.

**Conclusions:** Findings suggest that some young people may have felt less stigma talking about their mental health now compared to before the COVID-19 pandemic. However, many are worried about how the pandemic has affected their education and social connections and may require additional psychological, practical and social support. Our findings highlight the important role that education providers play in providing a source of information and support to adolescents and young adults during times of uncertainty.

## Introduction

When coronavirus disease 2019 (COVID-19) was declared a pandemic by the World Health Organisation (World Health Organisation, 2020) many governments imposed social distancing restrictions to suppress the virus, including self-isolation and mobility constraints. Whilst these measures are perceived to have been necessary to minimise contact and virus transmission, there is evidence to suggest that they have resulted in adverse psychological consequences across general populations (Niedzwiedz et al., 2020; Pierce et al., 2020).

Lessons from previous epidemics suggest that youth experience measures aimed at preventing the spread of viruses as particularly distressing and traumatic (Sprang & Silman, 2013) with the potential for some effects to be long-lasting, even after restrictions are lifted (Brooks et al., 2020; Kwong, Pearson, Smith, et al., 2020). Although children and adolescents are less likely to experience severe symptoms or hospitalisation with COVID-19 (Bhopal et al., 2021; Ludvigsson, 2020), the impact of quarantine represents a significant mental health threat (Fegert et al., 2020). Emerging evidence suggests that young people have experienced the greatest decline in mental health during the first wave of COVID-19, compared to all other age groups (Fancourt et al., 2020). Young people have reported increased depression (Barendse et al., 2021), anxiety (Kwong, Pearson, Adams, et al., 2020; Nearchou et al., 2020), uncertainty, grief, and frustration over government handling of the pandemic (Demkowicz et al., 2020).

Reasons for this reported decline in mental health among younger people are likely to be multifaceted. Firstly, school closures and education disruptions remove routine, structure and opportunities for socialisation (Lee, 2020), which may increase loneliness and isolation. For children or young people with mental health support needs, the closure of schools and universities removed access to coping resources or infrastructures located in these settings (e.g. mental health services, peer support or face-to-face services). Secondly, the age span from adolescence to young adulthood is a particularly sensitive one as young people experience major life transitions. Disruption to these transitions can cause uncertainty and anxiety, as documented in young adults experiencing job insecurity as a result of the pandemic (Ganson et al., 2021) or students experiencing the cancellation of their studies (Lee, 2020). Finally, beyond these individual factors, some adolescents and young adults faced additional stressors during COVID-19, including a decline in parental wellbeing, child maltreatment, and bereavement, all of which may compound any wider changes in circumstances initiated by the pandemic (e.g. online learning, home schooling) (Romanou & Belton, 2020; Young Minds, 2021). These experiences were likely felt most acutely by those already affected by socioeconomic marginalisation and deprivation (Waite et al., 2020).

In these contexts, young people and adolescents were at risk of unique psychosocial consequences from the pandemic compared to other population groups, and while others have sought to quantify the impact of the pandemic on young people’s mental health, there is limited work exploring young people’s individual perceptions. In this study, we aimed to explore adolescent and young adult experiences during the COVID-19 pandemic in the UK, specifically to understand how and why social distancing restrictions affected social lives, mental health, and wellbeing.

## Method

We undertook a qualitative study to explore the experiences of adolescents (aged 13-17) and young adults (aged 18-24) throughout the COVID-19 pandemic in the UK. This work forms part of the COVID-19 Social Study (CSS), where researchers are investigating the psychosocial wellbeing of people living in the UK during the pandemic (Bu, Steptoe, Mak, et al., 2020). The research was reviewed and approved by the University College London Ethics Committee (Project ID: 14895/005).

### Participant recruitment

We used a combination of convenience and purposive sampling strategies. Eligibility criteria included: aged 13-24, living in the UK, able to speak English sufficiently to understand the participant information and to participate in an interview. All participants aged 16-24 provided written informed consent. Participants aged 13-15 provided assent and a parent provided written informed consent. Those who returned a signed informed consent form were also asked to complete a demographics questionnaire (See Table 1 for participant demographics).

**Table 1:** Self-reported demographic characteristics

|  | Participants<br>n = 37 |
| --- | --- |
| **Age** |  |
| 13-14 | 4 |
| 15-16 | 12 |
| 17-18 | 8 |
| 19-20 | 2 |
| 21-22 | 6 |
| 23-24 | 5 |
| **Sex** |  |
| Female | 23 |
| Male | 14 |
| **Ethnicity** |  |
| White British | 27 |
| Mixed Race* | 5 |
| Asian and Asian British** | 4 |
| **Qualifications** |  |
| No qualifications | 9 |
| GSCE | 9 |
| A Levels | 13 |
| Undergraduate | 5 |
| Postgraduate | 1 |
| **Employment** |  |
| Still at school | 17 |
| At university | 14 |
| Fulltime employment | 3 |
| Apprenticeship | 1 |
| Self-employed | 1 |
| Unemployed | 1 |

\* Mixed Race – White and Black African, White and Black Caribbean, White and North African, Other Mixed Background. \*\*Asian or Asian British - Indian, Punjabi, Pakistani, Malaysian.

Participants were recruited via the CSS study newsletter, through partner organisations working with young people, personal contacts and for young adults aged 18+, via social media. All interested individuals were invited to contact the study team for further information. We screened those who registered their interest to ensure participants were from a range of diverse backgrounds in terms of age, ethnicity, and living situation. No participants were previously known to the interviewers prior to recruitment. Potential participants received a Study Information Sheet and all were given the opportunity to ask questions. All participants who participated in interviews were emailed an online £10 shopping voucher as a token of appreciation for their time.

### Data collection

All participant interviews took place remotely, either over telephone or video call. Experienced, postgraduate, male and female, qualitative health researchers conducted all interviews (AB, AM, JD, RC, SE, TM, LB). All interviewers have previously carried out interviews about mental health and social life during the COVID-19 pandemic for other CSS research work (Aughterson et al., 2021; Burton et al., 2020; May et al., 2021).

We conducted data collection as two separate groups, to ensure any newly identified themes discussed by participants in each age group were sufficiently explored during data collection and analysis. Interviews followed a semi-structured format (range: 17-65 minutes, average 38). For adolescents aged 13-17, interviews ranged from 17 to 46 minutes (average: 33 minutes) and for young adults aged 18-24 from 29 to 65 minutes (average: 44 minutes). As the study was designed to elicit experiences of social isolation and social restrictions on mental health, wellbeing and social lives, topic guide questions were informed by social network and sense of coherence theories (Antonovsky, 1979; Berkman, 1983). In brief, interview topics included: life prior to COVID-19, understanding of and adherence to social distancing guidelines, social lives, mental health, and prospection (for question examples, refer to Figure 1 and for the full topic guide, Appendix 1).

**Figure 1.**
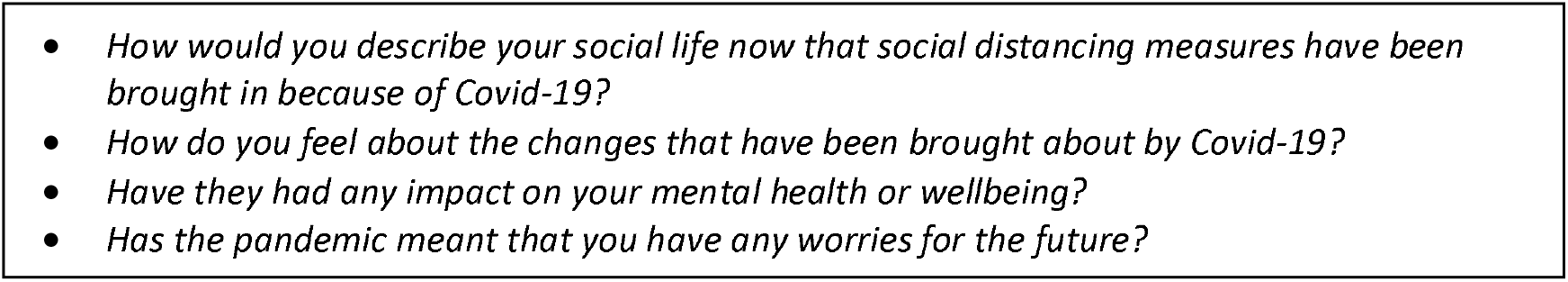
Interview Guide Examples

### Data analysis

All interviews were audio recorded and transcribed verbatim by an external transcription service. Returned transcripts were checked for anonymity and imported into NVivo version 12. We used a combination of inductive and deductive approaches during the analysis. First, an initial coding framework was developed based on the theoretical concepts explored in the interview topic guide. This was updated with new codes that were grounded in the data identified while reading through the transcripts. AM and TM then carried out line-by-line coding of interview transcripts and created the deductive coding framework. AM, TM and JD double coded three transcripts per group (adolescents, young adults) to ensure consistency in coding topics of salience. Finally, AM presented themes and subthemes during weekly internal meetings with the CSS research team to gain formative feedback on the comprehensibility of codes and preliminary findings.

## Results

We recruited 37 participants between June 2020 and January 2021. Participant characteristics are summarised in Table 1. The mean age of participants was 18 (range 13-24 years old). The majority were female (62%) and White British (73%). All participants lived with others (e.g., housemates, family, caregiver, spouse). Most participants were in secondary school or university (84%) and living at home with their parents (78%). Sixteen participants reported that they had an existing physical (7) and/or mental health condition (16).

We identified four overarching themes about the impact of the pandemic on mental health and wellbeing: a) Missing social contact during lockdown, b), Disruption to education, c) Changes to social relationships, and d) Improved wellbeing during lockdown. Themes and subthemes are illustrated in Figure 2.

**Figure 2:**
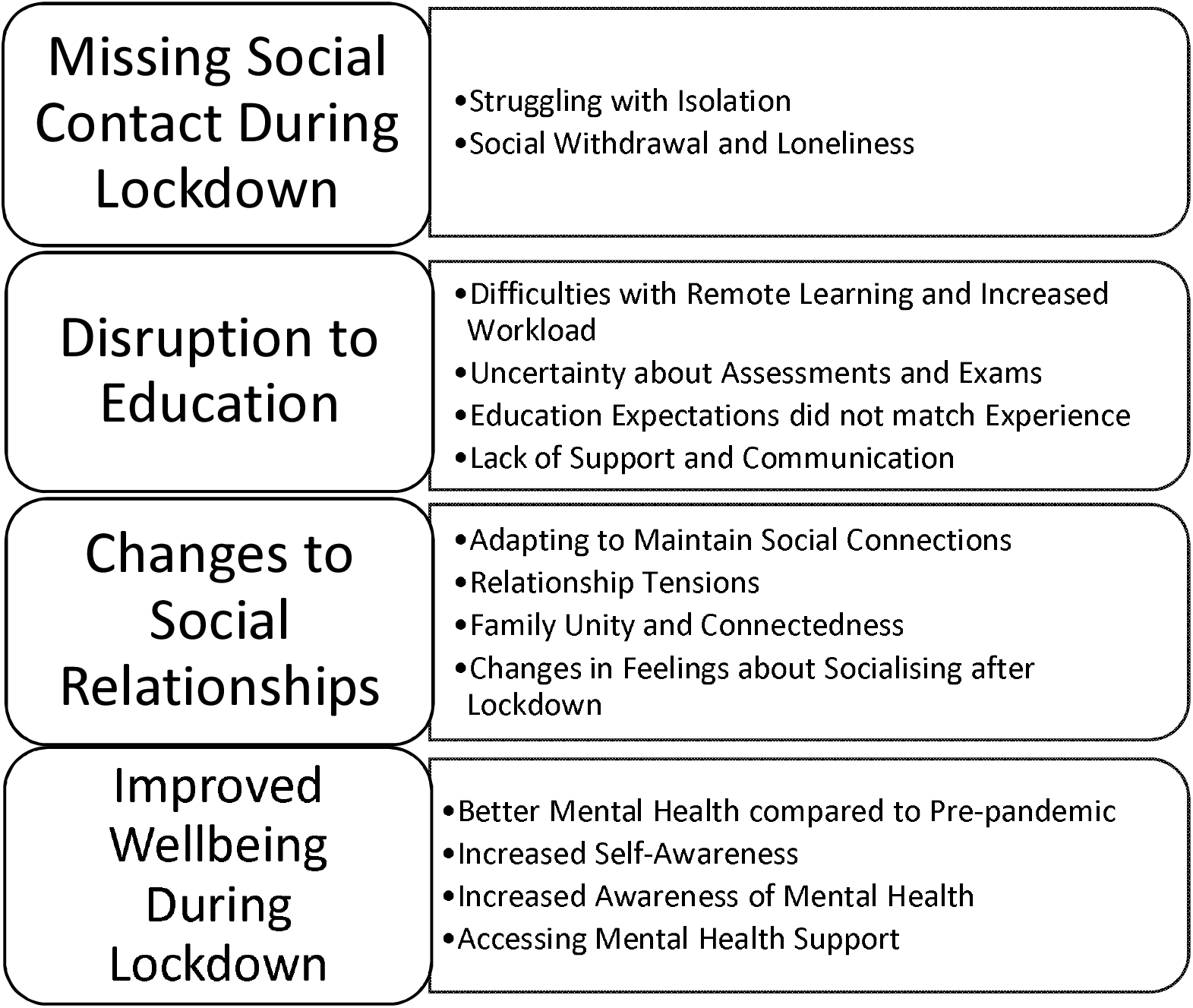
Themes and sub-themes

### Theme One: Missing Social Contact During Lockdown

Many participants reported a decline in wellbeing due to feeling lonely and isolated, particularly at the start of lockdown. Being unable to see friends or family and feeling that they were missing out on important life events was a source of sadness and frustration. Some withdrew themselves even further from online social activities after government restrictions were imposed.

#### Struggling with Isolation

Most participants said that their mental health was affected by social distancing restrictions. Many described this as being due to feeling socially restricted or “trapped” at home and being unable to maintain their usual activities and relationships.

> *“I’m not a very anxious person. I’ve struggled with mental health anyway, but… I think in the beginning of lockdown, being trapped at home wasn’t ideal in terms of my mental health*.*” P59, female, aged 18-22*

The feeling of being inside and at home for an indeterminate period of time was overwhelming for some, leading to a desire to overcompensate for what was perceived as “lost time” when restrictions began to ease after the first wave of the virus.

> *“As soon as restrictions were relaxed enough that you could meet someone socially distanced, I was down at my friends’ houses that are 100m down the road, three or four times a week, just to get out of my house. I got so frustrated being cooped up*.*” P55, female, aged 13-17*

The social distancing restrictions were especially challenging for participants who had pre-existing mental health conditions, who were more likely to report that their wellbeing had declined.

> *“…during lockdown, I got horrendously depressed. So, I’ve had stuff going on mentally, but I was finding it tough. The social isolation made everything crash really badly*.*” P39, male, aged 18-22*

### Social Withdrawal and Loneliness

While many participants had adapted their lives in response to social distancing restrictions, some, particularly adolescents, described socialising less during the lockdown and reported feelings of loneliness or social withdrawal.

> *“Most of my friends are in different countries… so, they were all in different time zones. So, I wasn’t able to speak to them much… It was so weird, especially because I’m an only child. So, it felt like I didn’t have anyone who I could really talk to who was around my age. So, it was really hard*.*” P51, female, aged 13-17*
>
> *“I don’t really speak to much of my mates as I used to. It’s made me quite distanced with quite a lot of people*.*” P66, female, aged 13-17*

At the start of the lockdown, some participants felt uncertain or overwhelmed by the move to online social interactions and made a conscious decision to withdraw from this method of contact.

> *“Before lockdown, I never used video calls or phoning people anyway. I spoke to people through messaging a lot, but I never used Zoom. I never heard of it to be honest. So, when I was suddenly forced to do that, it was just too much, so I’ve stopped talking to people*.*” P20, male, aged 13-17*

Boredom was commonly reported by a majority of participants and lowered their motivation to interact with others. Some participants said that they had nothing to talk about with their peers, or that friends were uninterested in keeping in contact. Therefore, efforts to maintain social connection ceased and usual social ties disrupted.

> *“I definitely tried to keep in touch with my school friends more at the start, but then it felt very one-sided, so I stopped making all the effort and I haven’t spoken to a few of them in a few months*.*” P44, female, aged 13-17*

Participants described feeling lonely throughout different stages of the pandemic, as though the world felt smaller from being at home during the lockdown without any social contact.

> *“There was some socialising over Zoom but overall, I think my world just sort of contracted fairly significantly…” P56, male, aged 18-22*

Several participants felt lonely even in the presence of others as restrictions began to ease, with many associating their feelings of loneliness with a lack of physical contact and closeness.

> *“So, even though we are back at school, we’re still struggling through the same things. We’re still feeling lonely, that we’re not able to have that physical contact, which I’m looking forward to having if the pandemic gets better*.*” P51, female, aged 13-17*

### Theme Two: Disruption to Education

The cancellation of assessments and closure of schools and universities were described as a source of loss, concern, and uncertainty for most participants. Many felt unsupported in their transition to online, remote learning and others were worried about the long-term impact of the pandemic on their prospects.

### Difficulties with Remote Learning and Increased Workload

Inability to concentrate, feelings of boredom and a lack of stimulation created difficulties with remote learning for some participants.

> *“I was just staring at a take home exam paper with my brain just completely blue screened. You know, just completely not functioning. I think that just general stimulus vacuum of lockdown massively contributed to that because that’s an experience I’ve never had before*.*” P56, male, aged 18-22*

For those living at home with their families, having to share a workspace with multiple siblings at once, or feeling confined to a bedroom to study because everyone in the household was working from home, meant that *“days felt like night*.*”*

> *“When I’ve sat in my room all day, there’s no distinguishing between the room where I sleep and the room where I do all my work*.*” P20, male, aged 13-17*

After several months of social distancing and online learning, some participants felt that the move to remote online learning had increased their workload compared to face-to-face learning.

> *“My school…said we’ll do live lessons, but then a teacher, instead of doing live lessons, just put the PowerPoint online, and then we just had to take notes from that. And they’d say, make four pages of notes on this, do the questions and then that’s your work done for the lesson, which wasn’t very good, because instead of spending one hour a lesson, I’d spend two or three hours*.*” P68, male, aged 13-17*

The impact of this increased workload on future exams and ultimately grades created an additional source of concern and uncertainty among participants.

> *“I know that the virus has definitely had a big toll on how much I’m thinking about [exams], because there’s so much uncertainty… My worries about exams [are] more like: ‘how am I going to get the content done in time,’ not how am I going to be graded*.*” P49, female, aged 13-17*

### Uncertainty about Assessments and Exams

Participants discussed feeling stressed, particularly at the start of lockdown, when there was a lack of clear guidance over how schools and universities would manage coursework and exams amid closures.

> *“It got announced the evening [school] closed there would be no GCSEs or A-Levels, but there was about a month on uncertainty of whether there would be more exams or anything similar. And that month was awful because we were still having to work but we were revising for stuff we knew we weren’t going to take. And teachers didn’t know, and we didn’t know, and there was so much uncertainty that it was awful*.*” P55, female, aged 13-17*
>
> *“University was probably biggest stress at the start because there was an uncertain period when we first went into lockdown, where all the universities were rushing to work out what they were going to do. At this point I was looking at handing in a dissertation and sitting three or four exams, and I didn’t know if it was still happening*.*” P54, female, aged 18-22*

When announcements were made that exams would be cancelled, many said that they felt upset or disappointed and reported feeling robbed of the chance to finish their education as planned.

> *“I’m actually really upset that I couldn’t sit my exams, so yes. It was a bit of an anticlimax, because I kind of wanted all of the build-up and the apprehension, to finalise, what, two years’ worth of… Well, my entire education built up to this. And when it was halted… I found it a bit difficult*.*” p59, female, aged 18-22*

Some participants reported that they had studied hard outside of school hours to improve their grades even before the pandemic hit and were anxious that this additional effort would not be taken into account when considering their grade predictions.

> *“Not doing exams, I was gutted. I can’t prove myself and I’m being given a letter that they think I could get*… *they didn’t see how hard I was working…they don’t see how hard we work outside of school. And even if, say, someone got a D but they pushed so hard trying to get an A and they were getting there, they were just trying so hard. They don’t see that, they don’t see everything we do in school. It’s just annoying that we can’t prove ourselves*.*” P34, female, aged 13-17*

### Education Expectations did not match Experience

Many participants spoke of feeling disappointment that their social plans for secondary school or university had not materialised due to closures and disruptions. Symbolic events tied to the start or end of education (such as “Freshers week”, “Leavers assembly” or prom) were cancelled and described as another source of loss during the pandemic.

> *“We didn’t get the finale of Year 13 like we normally do where there’s like a prom and things like that and it was all just a bit rushed so we didn’t get to say bye*.*” P53, male, aged 18-22*

Some said that they felt they had “missed out” on an experience that they saw as a rite of passage and one that they had been looking forward to.

> *“Because we were planning on going out so much and that all being cancelled, it was awful. The day when I was meant to have prom I put my prom dress on just because I wanted to… it was like the idea of it all being cancelled I felt like I was missing out*.*” P55, female, aged 13-17*

### Lack of Support and Communication

Many participants described feeling that there was a lack of support for their mental health and wellbeing from education providers during the pandemic.

> *“The school hasn’t been great in terms of providing help… the general attitude is: ‘it’s okay not to be okay, but we’re not going to do anything about it*.*’ I know a couple of other schools have a similar situation where you feel like, do they know that you’re struggling, but they’re not going to do anything*.*” P61, female, aged 13-17*

In cases where support was offered, some said that they had experienced this as performative rather than genuine.

> *“And I feel really let down by the school…Our head of year did a wellbeing check and it didn’t even seem genuine, it just seemed like she had to do it for her ego or something like that*…*” P34, female, aged 13-17*

Others described poorly managed communication and a lack of information from education providers about pandemic-related changes, which left students uncertain about their options for rearranging assessments and coursework.

> *“It would have been nice if I could speak to my personal tutor or something about it, but I think everyone was so busy at the beginning trying to record lectures and put them online, and everything was a bit hectic. So, if there was support it wasn’t very clear or easy to find,” P64, female, aged 18-22*

Poor communication and a perceived lack of support from education providers also resulted in some students feeling forgotten about or unfairly treated. This led to feelings of anger, frustration and a sense that their experiences were not valued.

> *“At the beginning, kind of March to May period, the response was absolutely shambolic and pretty malevolent to be honest…telling people to go home at their own expense up to and including sort of international students being told that they had to leave within a week even if that meant that their flights were two grand or something*.*” P56, male, aged 18-22*

### Theme Three: Changes to Social Relationships

#### Relationship Tensions

Participants spoke of stress, anxiety and anger regarding family and friendship tensions during the pandemic. For some, this was due to not being able to spend time with their friends, which led to relationship breakdowns.

> *“What’s changed is it’s put a lot of pressure on relationships, like with my boyfriend and a lot of people I know of have broken up with their boyfriends or girlfriends because of this quarantine and not being able to see them every day, or be around them as much*.*” P19, female, aged 13-17*

For others, differences in opinions between friends led to arguments over what was acceptable or not when following the social distancing rules.

> *“One of my mates, my mate, I actually ended up having an argument, because she was, like, oh, you’ve got to come and see me*.*” P66, female, aged 13-17*

Participants living with family members said that they experienced difficulties getting along with parents and siblings during lockdown, particularly when living together in small homes or when everyone was working from home.

> *“I don’t always get on with my family very well and we would be in each other’s faces a hell of a lot and that got very difficult at times… I’d say it gets worse when we have to spend more time together*.*” P38, male, aged 18-22*
>
> *“I think everyone just got a little bit stir crazy seeing the same walls and the same people*.*” P44, female, aged 13-17*

#### Relationship Unity and Connectedness

Despite relationship tensions, participants also described feeling settled and closer with their family members during the pandemic and lockdown periods.

> *“My relationship with my parents is great and it’s improved. It was at a great point this year and I think better than it’s ever been. And I think lockdown was a good time just to spend more time with them*.*” P37, female, aged 20-24*
>
> *“Obviously, my foster parents. And they keep me quite…Yes, they’re all right. They’ve been doing all right by me through this pandemic*.*” P66, female, aged 13-17*

Many were reminded about the importance of family and were resolute to make more effort to connect with others in the future.

> *“I won’t take for granted as much, like just standing outside talking to my neighbour, phone calls with my [relative]. Just the little things like that. Because obviously it’s so easy in normal life just to brush them aside and say, ‘oh, I’ll ring my [relative] later*.*’ So now I take the time to ring my [relative] and talk to her*.*” P33, female, aged 13-17*

Some participants had difficult conversations during the lockdown that they otherwise might not have had, for instance, what they would do if a family member became ill with the virus. Having frank and open conversations helped some to feel more connected than before the pandemic.

> *“I definitely think [the pandemic] forced us to talk about things that we wouldn’t always want to talk about. We had to discuss as a family what would happen to my dad if he was to get ill…We discussed those things already in general because we are quite open about that but I think it increased the intensity and frequency of those conversations about death and what would happen and health and things like that…those were conversations which I didn’t want to have with them so I think that, in a way, they were very positive and negative. But probably in the long run, they were positive because it’s helped us to be more open with each other” P41, female, aged 20-24*

#### Adapting to Maintain Social Connections

In response to social distancing measures, participants described the ways in which they had adapted their social lives, including trying new methods of online communication.

> *“The idea of not knowing when you’re next going to be in a crowded place, really able one on one to chat to people I think has just made people search for other ways of connecting with people and that’s online*.*” P41, female, aged 20-24*

Some said they were using dating apps, chatting through gaming platforms and sharing COVID-related memes as a way “to cope with difficult things” and enhance their sense of connectedness with others. New friendships were formed during lockdown with the help of group chats on gaming and social media platforms (i.e., “Whatsapp”).

> *“I have made a bunch of friends that are from online around my age from playing a game*…*so during the pandemic instead of chatting with my friends I’d be playing a game with them almost every day. Well, every day for a few hours chatting, having fun which was nice*.*” P21, male, aged 13-17*

#### Changes in Feelings about Socialising after Lockdown

Around half of participants described how their feelings about in-person social interactions had changed since the start of the pandemic. Some described this in terms of feeling more vigilant about being around other people and of the risks associated with future virus transmission.

> *“I think I’m going to be a lot more cautious about other people, obviously, like people you don’t know being quite close to you, physically*.*” P45, male, aged 13-17*
>
> *“I’m really apprehensive about the future of Covid, even though things are getting back to normal, and where we live things aren’t getting any worse, which is great. But it’s trusting other people, I think, which is quite a big thing, because you don’t know who everybody bothers with and stuff like that*.*” P33, female, aged 13-17*

Although individual concern about their own risk of catching COVID was not widespread in the group overall, some participants still reported feeling safer during lockdown away from other people and less exposed to the perceived risk of catching COVID-19.

> *“I felt less anxious about everything when we were in full lockdown and that the lifting out of lockdown, that’s quite scary in many ways for many different people…obviously introducing more risk into your life, whereas when everyone was in full lockdown, I feel like there was something quite comforting in a weird way about that*.*” P37, female, aged 20-24*

Adolescents in particular said they felt apprehensive about future social situations from not having been exposed to “real life” and becoming accustomed to life in lockdown.

> *“I just feel like I’ve lost the ability to form coherent sentences. I know that sounds really stupid. It’s just been really hard*.*” P34, female, aged 13-17*

Anxiety about socialising was especially present in those returning to school after months in lockdown, following difficulties in keeping in touch with friends and maintaining positive friendships.

> *“I was nervous, because a lot changed during quarantine. I think people got really tetchy staying at home, so there were a lot of arguments. Loads of people fell out of contact with one another…So, it was harder for us to find common things to talk about, we were just speaking about the same topics again and again. So, going back [to school, I] was really nervous because we didn’t know what we would talk about*.*” P49, female, aged 13-17*

### Theme Four: Improved Wellbeing During Lockdown

For those who described busy and stressful lives prior to the pandemic, social distancing restrictions provided a chance to pause and reflect, with resultant mental health benefits. Having more choice about how to spend free time was experienced by participants as relaxing and removed previous sources of stress. For some, their wellbeing was reportedly enhanced by support from others and an increased public awareness of mental health self-management during lockdown.

#### Improved Mental Health Compared to Pre-pandemic

In comparison to a majority of participants who reported struggling with their wellbeing during the pandemic, a smaller group of adolescents described how their mental health had improved during the first lockdown.

> *“Well, before lockdown, my mental health wasn’t great… my sleep schedule was all over the place, school stress, but actually, lockdown gave me a chance to focus and make myself a lot better in terms of the positive emotions” P49, female, aged 13-17*

They described busy or stressful school lives prior to the pandemic. School closures and a shift to online learning, particularly for those who were not preparing for their exams, meant that some had less work to do. The lockdown therefore provided an opportunity to relax and reflect.

> *“I think it’s been just I guess inconvenience rather than a full crisis in my eyes. It probably should have affected me more but, yes, I quite like it in quarantine to be honest. I don’t have to get up, I can just chill around. It’s a lot nicer than having to rush around constantly*.*” P21, male, aged 13-17*

As previously reported, the closure of schools and university meant an increased workload for some, but the flexibility of choosing what to do between lessons and during free time meant the lockdown felt “like a really long summer holiday” for others.

> *“I was happy when school got cancelled… it was still hard to do the work at home, but it was just because it was just like, in between I could just do whatever I wanted, so it was just better, sometimes I just go and play in the garden, football, or just be on my phone or the PS4*.*” p28, male, aged 13-17*

#### Increased Self-Awareness

Throughout the course of the pandemic, some participants had more time to take up hobbies and exercise, but several young adults also commented on the personal growth and increased self-awareness that they had observed.

> *“[This experience has] taught me to take everything a day as it comes. And then see what happens. But I won’t be surprised if, in a good way, that things change and it happens for the better*.*…it allowed me to challenge myself more*…*it allowed me to be better with talking to people. Because I could easily slur on words or I don’t know how to sum up anything. It just always used to be muddled up in my brain, whereas now things have started to flow better with how I am as a person*.*” P60, female, aged 18-22*

Some were reflecting on the stark changes in their lives during the pandemic compared with pre-pandemic and said they felt differently about how they wanted their life to be when social distancing restrictions eased.

> *“First, I think someone hit the brakes, I think, like everything stopped…[Life is] no longer that constant, hectic mess of going out…overall, it’s a lot less intense maybe, now*.*” P39, male, 18-22*

#### Increased Awareness of Mental Health

Some participants said they felt an increased awareness of their own mental health and how to protect it during the pandemic, through initiatives driven by workplaces and education providers.

> *“My organisation did allocate three weeks of wellbeing leave which is a really interesting move. And that was on top of annual leave. And that was complementary leave because the reason why they did it is that they said that it’s a really hard time and they just think everyone needs the break*.*” P37, female, aged 20-24*

Some said they had not paid attention to their mental health in the past compared to now.

> *“[the pandemic has] shown that health and mental health and mental state is important. As it always was, but I don’t think I realised it as much as I would now*.*” P46, male, aged 13-17*

This perceived change in mental health awareness helped make it feel easier to talk to others about their wellbeing, and for some, this “opened up” a dialogue with friends and family that had never been achieved before.

> *“It opened up really big conversations within our family about mental health that I don’t think they’ve ever had before like the generations. So those are two really big positives for me I think. Both myself and opening that up wider*…*It was just conversations that we’d never had before*.*” P37, female, aged 20-24*

### Accessing Mental Health Support

Some participants sought support to help cope with pandemic-related distress, either informally through friends and family, or formally through education providers, health services, or employers.

> *“I did talk to one of my teachers actually at school, on Teams…And she helped me and a lot of friends about things to do during lockdown*.*” P50, female, aged 13-17*
>
> *“So, first off I was talking to my friends and family almost every day… Like my really good friends and mum and dad all knew that I was really struggling because I obviously told them. So, they were checking in. I knew they were checking in regularly as well*.*” P37, female, aged 20-24*

In some cases, regular offers of support and check-ins by educational staff were sufficiently reassuring to know that help was on hand if needed.

> *“[University staff would] email every week, checking in. They gave me two… deadline extensions, for my work, which helped me a lot. And my tutor was really helpful. He did check in every couple of weeks to make sure I was all right. Even over the summer, when they haven’t necessarily been at uni. So, that was good*.*” P57, female, aged 18-22*

## Discussion

We sought the views of adolescents and young adults about their mental health and social lives during the COVID-19 pandemic using in-depth qualitative interviews. Young people described mental health consequences associated with lockdown measures, including loneliness, frustration, a sense of loss about cancelled plans and education disruptions, and uncertainty about socialising in the future. Our findings highlight how some felt paradoxically isolated but also hesitant about reintegrating with others as the lockdown restrictions ease. Despite the challenges, we also found evidence of a potentially positive impact of the pandemic, including an increased awareness of mental health self-management and having more open conversations about mental health compared to pre-pandemic.

Young people have reported some of the highest levels of loneliness during the COVID-19 pandemic (Bu, Steptoe, & Fancourt, 2020), which have been associated with a decline in mental health and wellbeing (Lee et al., 2020). Initiatives that enable young people to easily connect with others and access support for their mental health while adhering to social distancing guidelines are greatly needed (Lee et al., 2020). Participants in our study frequently cited mental health consequences due to social isolation and loneliness. These findings build on those shown in previous research involving young people during the first national lockdown (i.e., Demkowicz et al., 2020), including the sense of loss and uncertainty frequently reported by this group. Our results build on this by suggesting how social isolation led to these effects, including what felt like missed opportunities to create memories with friends and missed social events seen as rites of passage in young adulthood. Our results also shed light on the relationship between compliance and mental health in young people. Although amongst adults over 18, poor mental health was not found to predict lower adherence (Wright et al., 2021), a small number of participants in our study rationalised their non-adherence to guidelines as a means of protecting their mental health. This suggests that for younger age groups, protecting mental health was sometimes seen as equally or even more important than protecting themselves from catching COVID-19.

The effects of isolation and loneliness for young people are likely to persist for some time after the pandemic, even after restrictions have lifted (Loades et al., 2020). The path ahead will require additional resources for education providers and coordinated efforts from policy makers, education authorities, and health and social care to ensure that support and service provision is responsive to the needs of young people (Kim & Asbury, 2020). This response should be formulated in collaboration with young people to ensure that any future support incorporates appropriate coping strategies and protective activities (Efuribe et al., 2020), such as those identified by our study, including check-ins from education providers and wellbeing initiatives from employers. These responses could help to validate the unique struggles that many young people have faced throughout the different stages of the pandemic (Efuribe et al., 2020).

While the long-term effect of school closures on young people’s health, income and productivity is yet to be fully realised (Viner et al., 2020), disruptions to education on this scale have not occurred since the second World War (d’Orville, 2020). These changes will likely have long term consequences requiring ongoing support (The British Academy, 2021), with particular attention needed on how sociodemographic factors have affected young peoples’ experiences during the pandemic (Scott et al., 2021). Around 60% of students living in affluent areas had access to online learning during the pandemic compared with 23% of students living in more deprived areas (Cullinane & Montacute, 2020). A loss of learning has already been observed by teaching staff in the UK (Kim et al., 2021), and our findings suggest this may be compounded by increased workloads, sharing small workspaces at home, and difficulties with concentration. If unaddressed, this may restrict opportunities for employment and social mobility available to young people in the future (The British Academy, 2021). It is essential that those without the resources to work optimally from home are supported, to ensure educational and social inequalities are not exacerbated, both as COVID-19 persists and in other future scenarios where remote learning may be required.

Academic challenges and future worries have been frequently reported by young people during the pandemic, particularly among high-school aged youth (Scott et al., 2021). We also noted concern among those who had access to online learning but were unable to sit exams and finish their coursework. Several young people felt they did not demonstrate their full potential when their exams were cancelled and did not want to be known as the generation who were given a “free pass”. Teaching staff may be well-placed to address these concerns by clearly communicating revised assessment plans and providing reassurance about the assessment process for future students affected by pandemic-related education disruptions. Teachers also require clear guidance and coordination from government and educational authorities to be able to deliver these messages (Kim et al., 2021), which were absent during the first wave of educational establishment closures.

It is estimated more than half of students have experienced adverse mental health during the COVID-19 pandemic (Office for National Statistics, 2020). Based on the findings from this research and others (Marques de Miranda et al., 2020), young people are especially susceptible to mental health adversity during pandemics, particularly those experiencing hardship and inequality (Fegert et al., 2020; Rudenstine et al., 2021). We found that paradoxically, while some participants felt isolated and lonely, they were also apprehensive about social distancing restrictions easing and life returning to “normal”. Graded or phased approaches for those returning to education (Viner et al., 2021) and employment are recommended as lockdown restrictions ease (Habersaat et al., 2020), Shared spaces need to feel safe, hygienic and welcoming to assist those who are concerned with safety or experiencing concern about socialising again as a result of being isolated during the pandemic.

Despite the many negative effects of the pandemic on mental health in young people identified to date, evidence also suggests that adolescents’ prosocial behaviours have increased during the pandemic, including providing practical support to others and donating to charities in need (Alvis et al., 2020). It is encouraging that some participants in this study reported feeling more able to talk about their mental health compared with pre-pandemic. This was, at times, facilitated by top-down efforts from education providers, employers, and parents. In this study and others (Aughterson et al., 2021), we found that many have been reminded of the importance of family and friends during adversity. Some young people have planned to make positive changes in the way that they approach their relationships in response to their experience of living through COVID-19. These changes represent a potentially positive side effect of the pandemic, providing they are supported and maintained. Future interventions may therefore benefit from leveraging the support of others in attempts to buffer negative psychological consequences, given that so many young people found openly conversing with their social contacts about mental health to be beneficial.

### Strengths and Limitations

This is the first known UK study to use interview-based methods to learn more about the experiences of adolescents and young people during the COVID-19 pandemic. Purposive recruitment methods increased the variability of experiences within the group of participants interviewed. Adolescents and young adults were a range of ages, had different living situations and health statuses. Data were gathered throughout several key time points of the UK pandemic response, including as schools were reopening after the first lockdown, and as the second wave of the virus had begun. We relied on participants self-identifying to take part in the research, which may have resulted in a sample who were particularly motivated or whose experiences were more positive than those less comfortable or with less time to take part in research. Remote interviewing techniques may have excluded some groups with limited online access; however, we also offered to conduct telephone calls and remote data collection methods enabled us to speak to people from across the UK.

## Conclusions

When the pandemic began and restrictions were introduced, many young people lost a sense of certainty, community and belonging provided by their routines, social circles, and education providers. Our findings highlight how support for adolescents and young people to develop and maintain their social connections and access mental health support is needed even after the virus is controlled. Most young people in our study struggled with their mental health and loneliness during the pandemic, and our results highlight the vital role of education providers giving regular guidance and support. We also highlight the detrimental impact that an absence of information and support had on mental health. More must be done to ensure offers of support are genuine and reinforced with signposting to information, advice and services when young people do reach out for help.

## Data Availability

In order to preserve participant anonymity, research data are not available publically for this work.

## Acknowledgements

The researchers are grateful for the support of the British Youth Music Theatre during recruitment. Many thanks to Louise Baxter, Rana Conway and Sara Esser for their help with conducting interviews. Thank you to Katey Warren and Henry Aughterson who provided feedback on the themes and subthemes.

## Author Contribution

DF conceived the study and DF and AB designed the study. AB, AM, JD and TM collected data for the study, analysed and interpreted the data. AM wrote the first draft with suggestions from AB, JD and TM. All authors provided critical revisions, read, and approved the submitted manuscript. All authors had full access to the data in the study and can take responsibility for the integrity and accuracy of the data.

## Data Availability

The data are not publicly available due to their containing information that could compromise the privacy of research participants.

