## Appendix 1 for "“You’re just there, alone in your room with your thoughts” A qualitative study about the impact of lockdown among young people during the COVID-19 pandemic"

**Appendix 1: Topic Guide**

**Please could you describe what normal life was like for you before the pandemic and the social distancing? (can explain before – e.g. staying at home, stopping visiting people's houses, having to stand 2 metres apart, etc)**

- Who do you normally live with separated/ extended family?
- Do you have any siblings/ pets?
- School type, do you have friends you would normally visit or would visit you – please tell me about this?
- Do you have any hobbies or things you enjoyed doing out of school time?

UNDERSTANDING AND ADHERENCE TO GUIDELINES
**At the moment, are you 'staying at home'?**
- Can you tell me what this is like for you and your family?
    - i.e. are you/ your parents  going out for shopping or for walk/ exercise
    - Or staying inside with no outside exercise?/ other people dropping off shopping?

- **What do you understand by the 'social distancing' advice that is being given – what does it mean to you? (Ask to list the rules they are aware of)**
- Have you been…
    - Avoiding crowds
    - Keeping distance from others
    - Isolating (staying inside)
    - Avoiding close contact greetings
    - Socialising/going out only with those in your household

**Have you been able to stick to the social distancing advice that has been given? Please tell us about why/ why not?**
- What helps you to stick to it?
- Or – what are you finding difficult about sticking to the rules?
- What has motivated you to do this?
- Is there anything that makes it easier to do or harder

**[COM-B prompts can be used here, to include:]**
- Any existing physical or mental health problems
- Group membership/ applicability Whether it feels like the rules apply to you
- Beliefs about whether it will keep you healthy
- Beliefs about whether it will keep other people healthy
- Having to go out for things like food
- Work/ Caring responsibilities, providing emotional support
- Friends suggesting meeting, family or parents wanting to go out
- Government rules/punishments,
- Feelings about losing normal life
- Change of routine/ habits

**SOCIAL LIFE – In terms of before the pandemic, How would you describe your social life before the Covid-19 pandemic started?**
**Although some of the question seems less directly relevant, we are trying to understand a big picture of how groups affect/ influence how we feel.**

- How would you describe the group of people you knew before the pandemic (these may be the same as now)
  - Are they friends or family or both mainly?
  - Do they live near you or far away?
  - How often did you see each other?
- Did you speak to family and friends face to face/online?

- Were you involved in any groups outside school – please describe (examples such as choir, dancing classes, football teams, youth groups/ youth theatre, Scouts/Guides, social activities etc)
- Did you have people who helped you, such as teachers, Scout leaders, parents, grandparents, family friends? How did they help you, for example listening, help with homework, help with mental health?
- Can you tell us about whether or not your friendship groups encouraged you to get involved in things? Did you find you compared your life to theirs?
- Social roles possibly as a young carer/ older sibling

**How would you describe your social life now because of Covid-19?**

**Please tell us about this – make clear that we are talking about after the start of the pandemic, to see if anything has changed.**

Prompts include:

- How would you describe the group of people you know now?
  - Are they friends or family or both mainly?
  - Do they live near you or far away?
  - How often did you see each other?
- Do you speak to family and friends face to face/online?
- Are you seeing or speaking to teachers and students as part of school online?

- Are you involved in any online groups outside school – please describe (examples such as online choir, dancing classes, , youth groups/ youth theatre, Scouts/Guides, social activities etc)
- Do you have people who help you, such as teachers, Scout leaders, parents, grandparents, family friends? How did they help you, for example listening, help with homework, help with mental health?
- Can you tell us about any ways your friendship groups influence you such as encouraging you to get involved in things? Do you find you compare your life and theirs?
- Social roles possibly as a young carer

MENTAL HEALTH

**How do you feel about the changes that have been brought about by Covid-19?**
**Have they had any impact on your mental health or wellbeing? Please tell us about these**

- What are the things most bothering you at the moment?
- Have you experienced any impact on positive emotions? (prompts: concentration, being able to do and achieve things, relationships with others, how well you are managing and feelings of control over your situation?)
- Have you experienced any negative feelings? (prompts: such as, not enjoying things as much, anxiety, worry)
- Could you tell us if you have had any physical symptoms that might be due to being stressed or anxious? (prompts: fatigue, sleep problems, pain, illness symptoms, heart racing)

**Have you been doing/ planning anything to help with this?**

- Connecting with family or friends online
- Online groups?
- Hobbies/ Reading
- Exercise at home/ outside the home
- Volunteering
- Anything else you can tell me about?

**Why are you doing/ not doing these things?**

- Helpful/ not helpful – please tell us why
- Enjoyable
- Good for mental health/ wellbeing
- Can't get online, not connected, not comfortable, affordability, confidence in using/ skills
- Skills in using the internet/ communication software
- Living arrangements/ Work/ caring demands
- Peer support/ pressure
- Difficulties/ restriction in physical environment

PROSPECTION

**Has the pandemic meant that you have any worries for the future?**

**How are these different from the worries you had before?**

- Sense of control/ powerlessness
- Severity of worries / perspective

**Will this change the way you live your life in future?**

- The way you connect with others
- How you look after yourself
- How you support others
-

Has this changed what things are important to you?
